## Supplementary information for "Assessing and predicting adolescent and early adulthood common mental disorders in the ALSPAC cohort using electronic primary care data"

### Linkage to primary care data

The Secure Anonymised Information Linkage (SAIL) project has established a system which allows individual-level data from multiple sources – including electronic primary care data – to be linked together and analysed securely [1]. In collaboration with SAIL and GP software system providers, the Project to Enhance ALSPAC through Record Linkage (PEARL) developed two methods to extract primary care data. The first was a pilot extraction in 2012 using a sample of study participants based on an ‘opt-in’ consent principle, from which 2,806 participants consented to having their primary care records accessed (see [2] for further details). The second method was the main data extraction, where all ALSPAC participants with known addresses were sent fair processing materials and allowed to ‘opt-out’ of having their primary care records extracted, if they did not consent to this. Extraction of the electronic primary care data occurred in partnership with EMIS, which provides the software systems to most of the practices in the Bristol area. Records were extracted from all EMIS practices in the Bristol, North Somerset and South Gloucestershire clinical commissioning group which had consented to the NHS South West Commissioning Support Unit (SWCSU) governance framework and data extraction mechanism. Approval for this ALSPAC project was obtained from the SWCSU Security and Informatics Group and the SWCSU informed all participating practices and offered the option to opt-out. For more details, see [3].

Data processing methods after this extraction were identical for both the pilot and main extraction phases. The extracted electronic primary care records were pseudonymised and then securely transferred to a SAIL secure setting (a UK Secure eResearch Platform; UKSeRP) using SAIL’s ‘split file’ method. NHS standards of encryption and security were maintained throughout. These protocols have been described in more detail in previous publications [2,3].

*Table S1:* Read code lists

| Condition | Reference for Read code list (doi) / list of codes |
|---|---|
| **Anxiety – diagnosis, symptoms, treatment** | Cornish *et al.*, 2016: http://dx.doi.org/10.1136/bmjopen-2016-013167 |
| **Depression – diagnosis, symptoms, treatment** | Cornish *et al.*, 2016: http://dx.doi.org/10.1136/bmjopen-2016-013167 |
| **ADHD** | E2E.. (Childhood hyperkinetic syndrome)<br>E2E0. (Child attention deficit disorder)<br>E2E00 (Attention deficit without hyperactivity)<br>E2E01 (Attention deficit without hyperactivity)<br>E2E0z (Child attention deficit disorder NOS)<br>E2E1. (Hyperkinesis with developmental delay)<br>E2E2. (Hyperkinetic conduct disorder)<br>E2Ey. (Other hyperkinetic manifestation)<br>E2Ez. (Hyperkinetic syndrome NOS)<br>Eu90. ([X]Hyperkinetic disorders)<br>Eu900 ([X]Attention deficit hyperactivity disorder)<br>Eu901 ([X]Hyperkinetic conduct disorder)<br>Eu902 ([X]Deficits in attention motor control and perception)<br>Eu90y ([X]Other hyperkinetic disorders)<br>Eu90z ([X]Hyperkinetic disorder unspecified)<br>Eu9y7 ([X]Attention deficit disorder)<br>9Ngp. (On drug therapy ADHD)<br>8BPT. (Drug therapy for ADHD) |
| **Asthma** | Cornish *et al.*, 2014: http://dx.doi.org/10.1136/bmjopen-2014-005345 |
| **Autism Spectrum Disorder (ASD)** | Hagberg & Jick, 2017: https://doi.org/10.2147/CLEP.S139107 |
| **Bipolar disorder: diagnosis** | Carr *et al.*, 2017: https://dx.doi.org/10.1370%2Fafm.2054 (used diagnosis codes only from their list) |
| **Conduct disorder** | E2C4. (Mixed disturbance of conduct and emotion)<br>E2C4z (Mixed disturbance of conduct and emotion NOS)<br>Eu912 ([X]Socialized conduct disorder)<br>Eu911 ([X]Conduct disorder, solitary aggressive type)<br>E2C2z (Socialised conduct disorder NOS)<br>E2C1. (Nonaggressive unsocial conduct disorder)<br>Eu911 ([X]Unsocialized conduct disorder)<br>E2C.. (Disturbance of conduct NEC)<br>Eu91. ([X]Conduct disorders)<br>Eu92z ([X]Mixed disorder of conduct and emotions, unspecified)<br>Eu92. ([X]Mixed disorders of conduct and emotions)<br>E2C2. (Socialised conduct disorder)<br>E2C0. (Aggressive unsocial conduct disorder)<br>E2C1z (Nonaggressive unsocial conduct disorder NOS)<br>E2C0z (Aggressive unsocial conduct disorder NOS)<br>Eu901 ([X]Hyperkinetic disorder associated with conduct disorder)<br>E2Czz (Disturbance of conduct NOS)<br>Eu91z ([X]Childhood conduct disorder NOS)<br>Eu92y ([X]Conduct disorder associated with emotional disorder)<br>Eu91y ([X]Other conduct disorders)<br>E2Cy. (Other conduct disturbances)<br>E2Cz. (Unspecified disturbance of conduct) |

| | |
|---|---|
| | E2Cyz (Other conduct disturbances NOS)<br>Eu910 ([X]Conduct disorder confined to the family context)<br>Eu91z ([X]Conduct disorder, unspecified)<br>Eu920 ([X]Depressive conduct disorder)<br>Eu92y ([X]Other mixed disorders of conduct and emotions)<br>Eu913 ([X]Oppositional defiant disorder)<br>E2Dy0 (Childhood and adolescent oppositional disorder) |
| **Eating disorder** | Carr *et al*., 2017: https://dx.doi.org/10.1370%2Fafm.2054 |
| **Eczema** | Mukherjee *et al*., 2014: https://dx.doi.org/10.1136/bmjopen-2014-006647 (used diagnosis codes only from their list) |
| **Family history depression** | Read codes 1285. |
| **Family history mental illness** | 128% except 1281. (Read codes starting 128 - % used as sort of wild card for Read codes. 1281. is family history senile dementia) |
| **General symptoms** | 1954. (Indigestion)<br>1968. (Abdominal discomfort)<br>1969. (Abdominal pain)<br>1981. (c/o Nausea)<br>1982. (Nausea present)<br>198Z. (Nausea NOS)<br>1B14. (Tenseness)<br>1B18. (Inadequate)<br>1B1G. (Headache)<br>1B1G1 (c/o Headache)<br>1B1N. (Poor self-esteem)<br>1B1O. (Restless)<br>1B51 (Dizziness symptoms)<br>E2781 (Tension headache)<br>R0040 ([D] Dizziness)<br>R00Z2 ([D] Pain, generalised)<br>R040. ([D] Headache)<br>R0700 ([D] Nausea)<br>R090. ([D] Abdominal pain)<br>R090z ([D] Abdominal pain NOS)<br>R090E ([D] Recurrent abdominal pain)<br>R090N ([D] Non-specific abdominal pain) |
| **History of depression** | 1465. (H/O: depression) |
| **History of mental illness** | 146%. (H/O: psychiatric disorder) |
| **Number of consultations and prescriptions** | Cornish *et al*., 2020: https://doi.org/10.1093/ije/dyaa192 |
| **Psychosis** | Carr *et al*., 2017: https://dx.doi.org/10.1370%2Fafm.2054 (used diagnosis codes only from their list) |
| **Referral to mental health services** | 8H49. (Psychiatric referral)<br>8H4P. (Referral to child psychiatrist)<br>8H78. (Referral to counsellor)<br>8H7A. (Referral to mental health worker)<br>8H7B. (Referral to community psychiatric nurse)<br>8H7T. (Referral to psychotherapist)<br>8H7W. (Referral to TOP counselling)<br>8Hc.. (Referral to mental health team)<br>8Hc0. (Referral to community mental health team)<br>8Hc1. (Referral to mental health crisis team) |

| | |
|---|---|
| | 8Hc2. (Referral to primary care mental health team)<br>8HHn. (Referral to non-NHS mental health community service)<br>8HHp. (Referral for guided self-help for anxiety)<br>8HHq. (Referral for guided self-help for depression)<br>8HHr. (Referral for mental health self-help literature)<br>8HHu. (Referral to primary care mental health gateway worker)<br>8HHR. (Referral to child and adolescent psychiatry service)<br>8HHT. (Refer to psychotherapist)<br>8HkK. (Referral to improving access to psychological therapies programme)<br>8HVO. (Private referral to psychiatrist) |
| **Schizophrenia** | Carr *et al*., 2017: https://dx.doi.org/10.1370%2Fafm.2054 (used diagnosis codes only from their list) |
| **Self-harm** | Thomas *et al*., 2012 : https://dx.doi.org/10.1111%2Fbcp.12059 |
| **Smoking** | Cornish *et al*., 2020: https://doi.org/10.1093/ije/dyaa192 |
| **Somatic symptoms** | 16122 (Loss of appetite – symptom)<br>1613. (Increased appetite)<br>1615. (Reduced appetite)<br>168% (Tired all the time, fatigue, tiredness, tearful, malaise/lethargy)<br>1B15. (Irritable)<br>1B16. (Agitated)<br>1B19. (Suicidal)<br>1B1B. (Cannot sleep – insomnia)<br>1B1D. (Nightmares)<br>1B1I. (Crying, excessive)<br>1B1I1 (C/O tearfulness)<br>1BD1. (Suicidal ideation)<br>1BD2. (Morbid thoughts)<br>1BD3. (Suicidal plans)<br>1BD4. (Suicide risk)<br>1BD5. (High suicide risk)<br>1BD6. (Moderate suicide risk)<br>1BD8. (At risk of deliberate self harm)<br>1BDA. (Thoughts of deliberate self harm)<br>1BJ.. (Loss of confidence)<br>1BR.. (Reduced concentration)<br>1BW.. (Poor concentration)<br>E2052 (Tired all the time)<br>E2741 (Insomnia NOS) |
| **Substance abuse (alc/drugs)** | Eu1% ([X] Mental and behavioural disorders due to psychoactive substance abuse)<br>E23% (Alcohol dependence)<br>E24% (Drug dependence) |

*Table S2:* Details of variables used in the misclassification analyses.

| Variable | Origin | Type | Notes |
|---|---|---|---|
| **Sex** | ALSPAC | Binary | 0 = Male; 1 = Female |
| **Age at data collection event** | ALSPAC | Cont. | Measured in months |
| **Mother's age at birth** | ALSPAC | Cont. | Measured in years |
| **Mother's home ownership status** | ALSPAC | Binary | 0 = Owned/Mortgaged; 1 = Rented/Housing Association/Council House/Other |
| **Mother's marital status** | ALSPAC | Binary | 0 = Married; 1 = Not married |
| **Mother's age at first pregnancy** | ALSPAC | Cont. | Measured in years |
| **Mother's parity at child's birth** | ALSPAC | Binary | 0 = First child; 1 = Second or later child |
| **Mother's highest education level** | ALSPAC | Binary | 0 = CSE/Vocational/O Level; 1 = A Level/Degree |
| **Father's highest education level** | ALSPAC | Binary | 0 = CSE/Vocational/O Level; 1 = A Level/Degree<br>Assumes Mother's partner is child's Father. |
| **Mother's social class** | ALSPAC | Binary | 0 = I/II – Professional/Managerial; 1 – III/IV/V – Skilled/Partly skilled/Unskilled |
| **Father's social class** | ALSPAC | Binary | 0 = I/II – Professional/Managerial; 1 – III/IV/V – Skilled/Partly skilled/Unskilled<br>Assumes Mother's partner is child's Father. |
| **Child's ethnic background** | ALSPAC | Binary | 1 = White; 2 = Non-white |
| **Child's education (# A*-C GCSE equivalents)** | ALSPAC | Cont. | From education linkage data |
| **CIS-R depression score** | ALSPAC | Cat. | Specific to TF4 analyses |
| **CIS-R diagnosis of moderate depressive episode** | ALSPAC | Binary | Specific to TF4 analysis |
| **CIS-R diagnosis of severe depressive episode** | ALSPAC | Binary | Specific to TF4 analysis |
| **CIS-R Generalised Anxiety Disorder (GAD) symptoms** | ALSPAC | Binary | Specific to TF4 analysis |
| **CIS-R total score** | ALSPAC | Cont. | Specific to TF4 analysis |
| **DAWBA predicted depression** | ALSPAC | Cat. | Specific to TF3 analyses |
| **DAWBA predicted anxiety** | ALSPAC | Cat. | Specific to TF3 analyses |
| **Total MFQ score** | ALSPAC | Cont. | Specific to CCS, CCT, YPA and YPB analyses |
| **Historical depression diagnosis** | Primary care | Binary | 0 = No; 1 = Yes |
| **Historical anxiety diagnosis** | Primary care | Binary | 0 = No; 1 = Yes |
| **Current anxiety diagnosis** | Primary care | Binary | 0 = No; 1 = Yes |
| **Historical phobia diagnosis** | Primary care | Binary | 0 = No; 1 = Yes |
| **Current phobia diagnosis** | Primary care | Binary | 0 = No; 1 = Yes |
| **Historical depression symptoms** | Primary care | Binary | 0 = No; 1 = Yes |
| **Current depression symptoms** | Primary care | Binary | 0 = No; 1 = Yes |
| **Historical anxiety symptoms** | Primary care | Binary | 0 = No; 1 = Yes |
| **Current anxiety symptoms** | Primary care | Binary | 0 = No; 1 = Yes |

| | | | |
|---|---|---|---|
| **Historical somatic symptoms** | Primary care | Binary | 0 = No; 1 = Yes |
| **Current somatic symptoms** | Primary care | Binary | 0 = No; 1 = Yes |
| **Historical general symptoms** | Primary care | Binary | 0 = No; 1 = Yes |
| **Current general symptoms** | Primary care | Binary | 0 = No; 1 = Yes |
| **Historical anti-depressant treatment** | Primary care | Binary | 0 = No; 1 = Yes |
| **Current anti-depressant treatment** | Primary care | Binary | 0 = No; 1 = Yes |
| **Historical anti-anxiety treatment** | Primary care | Binary | 0 = No; 1 = Yes |
| **Current anti-anxiety treatment** | Primary care | Binary | 0 = No; 1 = Yes |
| **# of GP consultations per year** | Primary care | Cont. | Average over 2-year period at time of data collection |
| **# of GP prescriptions per year** | Primary care | Cont. | Average over 2-year period at time of data collection |
| **Asthma diagnosis** | Primary care | Binary | 0 = No; 1 = Yes (Includes any previous read codes, up to 6 months after age at event) |
| **Eczema diagnosis** | Primary care | Binary | 0 = No; 1 = Yes (Includes any previous read codes, up to 6 months after age at event) |
| **Eating disorder** | Primary care | Binary | 0 = No; 1 = Yes (Includes any previous read codes, up to 6 months after age at event) |
| **ADHD** | Primary care | Binary | 0 = No; 1 = Yes (Includes any previous read codes, up to 6 months after age at event) |
| **Conduct disorder** | Primary care | Binary | 0 = No; 1 = Yes (Includes any previous read codes, up to 6 months after age at event) |
| **Autism spectrum disorder** | Primary care | Binary | 0 = No; 1 = Yes (Includes any previous read codes, up to 6 months after age at event) |
| **Substance abuse (alcohol/drugs)** | Primary care | Binary | 0 = No; 1 = Yes (Includes any previous read codes, up to 6 months after age at event) |
| **Family history of mental illness** | Primary care | Binary | 0 = No; 1 = Yes (Includes any previous read codes, up to 6 months after age at event) |
| **Family history of depression** | Primary care | Binary | 0 = No; 1 = Yes (Includes any previous read codes, up to 6 months after age at event) |
| **History of mental health issues** | Primary care | Binary | 0 = No; 1 = Yes (Includes any previous read codes, up to 6 months after age at event) |
| **History of depression** | Primary care | Binary | 0 = No; 1 = Yes (Includes any previous read codes, up to 6 months after age at event) |
| **Self-harm** | Primary care | Binary | 0 = No; 1 = Yes (Includes any previous read codes, up to 6 months after age at event) |
| **Other psychological illness (schizophrenia, bipolar disorder and psychosis)** | Primary care | Binary | 0 = No; 1 = Yes (Includes any previous read codes, up to 6 months after age at event) |
| **Any other mental health issues** | Primary care | Binary | 0 = No; 1 = Yes (Combining eating disorder, ADHD, conduct disorder, ASD, substance abuse and other psychological illness, as low cell counts for these individual variables) |
| **Referral to mental health services** | Primary care | Binary | 0 = No; 1 = Yes (Includes any previous read codes, up to 6 months after age at event) |
| **Smoking (or ex-smoker)** | Primary care | Binary | 0 = No; 1 = Yes (Includes any previous read codes, up to 6 months after age at event) |

*Table S3:* Details of variables used in the lasso analyses.

| Variable | Type | Notes |
|---|---|---|
| **Sex** | Binary | 0 = Male; 1 = Female |
| **Age at ALSPAC data collection** | Cont. | Measured in months |
| **Historical depression diagnosis** | Binary | 0 = No; 1 = Yes |
| **Historical anxiety diagnosis** | Binary | 0 = No; 1 = Yes |
| **Current anxiety diagnosis** | Binary | 0 = No; 1 = Yes |
| **Historical phobia diagnosis** | Binary | 0 = No; 1 = Yes |
| **Current phobia diagnosis** | Binary | 0 = No; 1 = Yes |
| **Historical depression symptoms** | Binary | 0 = No; 1 = Yes |
| **Current depression symptoms** | Binary | 0 = No; 1 = Yes |
| **Historical anxiety symptoms** | Binary | 0 = No; 1 = Yes |
| **Current anxiety symptoms** | Binary | 0 = No; 1 = Yes |
| **Historical somatic symptoms** | Binary | 0 = No; 1 = Yes |
| **Current somatic symptoms** | Binary | 0 = No; 1 = Yes |
| **Historical general symptoms** | Binary | 0 = No; 1 = Yes |
| **Current general symptoms** | Binary | 0 = No; 1 = Yes |
| **Historical anti-depressant treatment** | Binary | 0 = No; 1 = Yes |
| **Current anti-depressant treatment** | Binary | 0 = No; 1 = Yes |
| **Historical anti-anxiety treatment** | Binary | 0 = No; 1 = Yes |
| **Current anti-anxiety treatment** | Binary | 0 = No; 1 = Yes |
| **# of GP consultations per year** | Cont. | Average over 2-year period at time of data collection |
| **# of GP prescriptions per year** | Cont. | Average over 2-year period at time of data collection |
| **Asthma diagnosis** | Binary | 0 = No; 1 = Yes (Includes any previous read codes, up to 6 months after age at event) |
| **Eczema diagnosis** | Binary | 0 = No; 1 = Yes (Includes any previous read codes, up to 6 months after age at event) |
| **Eating disorder** | Binary | 0 = No; 1 = Yes (Includes any previous read codes, up to 6 months after age at event) |
| **ADHD** | Binary | 0 = No; 1 = Yes (Includes any previous read codes, up to 6 months after age at event) |
| **Conduct disorder** | Binary | 0 = No; 1 = Yes (Includes any previous read codes, up to 6 months after age at event) |
| **Autism spectrum disorder** | Binary | 0 = No; 1 = Yes (Includes any previous read codes, up to 6 months after age at event) |
| **Substance abuse (alcohol/drugs)** | Binary | 0 = No; 1 = Yes (Includes any previous read codes, up to 6 months after age at event) |
| **Family history of mental illness** | Binary | 0 = No; 1 = Yes (Includes any previous read codes, up to 6 months after age at event) |
| **Family history of depression** | Binary | 0 = No; 1 = Yes (Includes any previous read codes, up to 6 months after age at event) |

| | | |
|---|---|---|
| **History of mental health issues** | Binary | 0 = No; 1 = Yes (Includes any previous read codes, up to 6 months after age at event) |
| **History of depression** | Binary | 0 = No; 1 = Yes (Includes any previous read codes, up to 6 months after age at event) |
| **Self-harm** | Binary | 0 = No; 1 = Yes (Includes any previous read codes, up to 6 months after age at event) |
| **Other psychological illness (schizophrenia, bipolar disorder and psychosis)** | Binary | 0 = No; 1 = Yes (Includes any previous read codes, up to 6 months after age at event) |
| **Any other mental health issues** | Binary | 0 = No; 1 = Yes (Combining eating disorder, ADHD, conduct disorder, ASD, substance abuse and other psychological illness, as low cell counts for these individual variables) |
| **Referral to mental health services** | Binary | 0 = No; 1 = Yes (Includes any previous read codes, up to 6 months after age at event) |
| **Smoking (or ex-smoker)** | Binary | 0 = No; 1 = Yes (Includes any previous read codes, up to 6 months after age at event) |

*Table S4:* Reasons for having ALSPAC clinic/questionnaire common mental disorder (CMD) data, but not primary care linkage data. Note also that the numbers excluded do not always correspond to the numbers with ALSPAC, but not primary care, data; this is because some individuals may have been counted both in the 'lost from records' and 'first entered records' columns (i.e., they first have primary care data within 18 months of the time-point, then were lost from primary care records again within 6 months after the time-point), or have missing data. Numbers in brackets denote the percentage of individuals in each exclusion category of the total participants without primary care data but with ALSPAC data. Most individuals are likely to be lost from primary care records because they moved out of the Bristol area, but it is also possible that they stayed within the Bristol area but moved to a GP practice using a different software system from which it was not possible to extract primary care data. Similarly, individuals may first appear in the primary care data after the time-point because they either moved into the Bristol area at this time, or stayed in the Bristol area but moved to a GP practice from which it was possible to extract primary care data.

| **Age (time point)** | **# with ALSPAC CMD data** | **# with ALSPAC, but not primary care, CMD data** | **# Excluded as no primary care data (no consent or no records)** | **# Excluded as lost from primary care records before time-point** | **# Excluded as first primary care data is after time-point** |
|---|---|---|---|---|---|
| **Age 15/16 (TF3 clinic)** | 5,332 | 1,669 | 737 (44.2%) | 614 (36.8%) | 313 (18.8%) |
| **Age 16/17 (CCS quest)** | 4,950 | 1,737 | 762 (43.9%) | 690 (39.7%) | 283 (16.3%) |
| **Age 17/18 (TF4 clinic)** | 4,534 | 1,450 | 583 (40.2%) | 616 (42.5%) | 254 (17.5%) |
| **Age 18/19 (CCT quest)** | 3,302 | 1,320 | 510 (38.6%) | 601 (45.5%) | 214 (16.2%) |
| **Age 21/22 (YPA quest)** | 3,283 | 1,985 | 471 (23.7%) | 1,462 (73.7%) | 64 (3.2%) |
| **Age 22/23 (YPB quest)** | 3,896 | 2,571 | 563 (21.9%) | 1,969 (76.6%) | 43 (1.7%) |

*Table S5:* Demographics at age 16/17 CCS questionnaire, 17/18 TF4 clinic, 18/19 CCT questionnaire and 21/22 YPA questionnaire time points. Of those with ALSPAC data, the table compares those who have primary data against those who do not. For categorical variables cells are counts and percentages. For continuous variables cells are means and standard deviations. Note also that the denominators vary as the variables come from different data sources, with different levels of completeness. See table 3 for age 15/16 TF3 and 22/23 YPB questionnaire time points.

| | **Age 16/17 (CCS questionnaire) – SMFQ (Depression)** | | **Age 17/18 (TF4 clinic) – CIS-R (CMDs and Depression)** | | **Age 18/19 (CCT questionnaire) – SMFQ (Depression)** | | **Age 21/22 (YPA questionnaire) – SMFQ (Depression)** | |
|---|---|---|---|---|---|---|---|---|
| | **Primary care data (*n*=3213)** | **No primary care data (*n*=1737)** | **Primary care data (*n*=3084)** | **No primary care data (*n*=1450)** | **Primary care data (*n*=1982)** | **No primary care data (*n*=1320)** | **Primary care data (*n*=1228)** | **No primary care data (*n*=1985)** |
| **Sex** | | | | | | | | |
| **Male** | 1302 (40.5%) | 706 (40.6%) | 1358 (44.0%) | 624 (43.0%) | 703 (35.5%) | 470 (35.6%) | 456 (35.1%) | 710 (35.8%) |
| **Female** | 1911 (59.5%) | 1031 (59.4%) | 1726 (56.0%) | 826 (57.0%) | 1279 (64.5%) | 850 (64.4%) | 842 (64.9%) | 1275 (64.2%) |
| **Maternal age at child's birth** | 29.4 (4.6) | 29.4 (4.5) | 29.3 (4.6) | 29.2 (4.6) | 29.4 (4.6) | 29.8 (4.6) | 29.2 (4.6) | 29.8 (4.4) |
| **Mother's home ownership status** | | | | | | | | |
| **Owned/ Mortgaged** | 2549 (85.8%) | 1359 (83.6%) | 2405 (85.4%) | 1115 (83.7%) | 1567 (85.2%) | 1052 (85.1%) | 1015 (85%) | 1632 (87.6%) |
| **Rented** | 138 (4.6%) | 104 (6.4%) | 113 (4%) | 69 (5.2%) | 83 (4.5%) | 70 (5.7%) | 53 (4.4%) | 94 (5%) |
| **Council/Housing Association** | 220 (7.4%) | 101 (6.2%) | 229 (8.1%) | 101 (7.6%) | 148 (8.1%) | 72 (5.8%) | 98 (8.2%) | 89 (4.8%) |
| **Other** | 65 (2.2%) | 62 (3.8%) | 69 (2.5%) | 48 (3.6%) | 41 (2.2%) | 42 (3.4%) | 28 (2.4%) | 49 (2.6%) |
| **Mother's marital status** | | | | | | | | |
| **Never married** | 380 (12.7%) | 214 (13%) | 386 (13.6%) | 190 (14.1%) | 242 (13%) | 130 (10.4%) | 146 (12.1%) | 206 (11%) |
| **Single/Divorced** | 132 (4.4%) | 86 (5.2%) | 122 (4.3%) | 70 (5.2%) | 78 (4.2%) | 56 (4.5%) | 48 (4%) | 80 (4.3%) |
| **First marriage** | 2305 (76.9%) | 1225 (74.6%) | 2165 (76.3%) | 998 (74%) | 1435 (77%) | 979 (78.2%) | 924 (76.6%) | 1470 (78.4%) |
| **$2^{nd}/3^{rd}$ marriage** | 182 (6.1%) | 118 (7.2%) | 164 (5.8%) | 91 (6.8%) | 108 (5.8%) | 87 (7%) | 88 (7.3%) | 120 (6.4%) |
| **Mother's parity** | | | | | | | | |
| **0** | 1410 (47.3%) | 840 (51.5%) | 1326 (47.2%) | 670 (50.6%) | 876 (47.4%) | 629 (50.8%) | 556 (46.5%) | 933 (50.2%) |
| **1** | 1058 (35.5%) | 553 (33.9%) | 999 (35.6%) | 444 (33.6%) | 666 (36%) | 413 (33.4%) | 433 (36.2%) | 631 (34%) |
| **2 or more** | 511 (17.5%) | 238 (14.6%) | 482 (17.2%) | 209 (15.8%) | 307 (16.6%) | 196 (15.8%) | 207 (17.3%) | 294 (15.8%) |
| **Mother's highest education level** | | | | | | | | |
| **O level/lower** | 1612 (54.1%) | 777 (47.7%) | 1541 (55.2%) | 642 (48.2%) | 993 (53.9%) | 542 (43.6%) | 726 (60.8%) | 826 (44.3%) |
| **A level** | 825 (27.7%) | 468 (28.7%) | 756 (27.1%) | 400 (30%) | 496 (26.9%) | 367 (29.6%) | 300 (25.1%) | 555 (29.7%) |
| **Degree** | 544 (18.3%) | 385 (23.6%) | 495 (17.7%) | 291 (21.8%) | 355 (19.3%) | 333 (26.8%) | 168 (14.1%) | 485 |

| | | | | | | | | |
|---|---|---|---|---|---|---|---|---|
| | | | | | | | | (26%) |
| **Father's highest education level** | | | | | | | | |
| **O level/lower** | 1217 (44.4%) | 556 (36.6%) | 1143 (44.5%) | 502 (40.7%) | 737 (43.7%) | 391 (33.2%) | 538 (49.3%) | 604 (34.2%) |
| **A level** | 828 (30.2%) | 465 (30.6%) | 795 (30.9%) | 363 (29.4%) | 510 (30.3%) | 365 (31%) | 344 (31.5%) | 528 (29.9%) |
| **Degree** | 699 (25.5%) | 498 (32.8%) | 632 (24.6%) | 368 (29.9%) | 439 (26%) | 423 (35.9%) | 209 (19.2%) | 634 (35.9%) |
| **Child ethnicity** | | | | | | | | |
| **White** | 2836 (96.4%) | 1535 (95.6%) | 2639 (95.8%) | 1255 (95.4%) | 1752 (96%) | 1186 (96.3%) | 1138 (96.2%) | 1769 (95.9%) |
| **Non-white** | 105 (3.6%) | 70 (4.4%) | 116 (4.2%) | 60 (4.6%) | 74 (4%) | 46 (3.7%) | 45 (3.8%) | 75 (4.1%) |
| **# GCSEs (or equivalents)** | 7.7 (3.4) | 7.8 (3.4) | 7.5 (3.5) | 7.6 (3.4) | 7.8 (3.3) | 8.4 (3.1) | 7.5 (3.5) | 8.5 (3) |
| **ALSPAC depression diagnosis** | | | | | | | | |
| **No** | 2728 (84.9%) | 1452 (83.6%) | 2841 (92.1%) | 1334 (92%) | 1595 (80.5%) | 1095 (83%) | 1098 (84.6%) | 1685 (84.9%) |
| **Yes** | 485 (15.1%) | 285 (16.4%) | 243 (7.9%) | 116 (8%) | 387 (19.5%) | 225 (17.1%) | 200 (15.4%) | 300 (15.1%) |
| **ALSPAC common mental disorder (CMD) diagnosis** | | | | | | | | |
| **No** | - | - | 2629 (85.3%) | 1231 (84.9%) | - | - | - | - |
| **Yes** | - | - | 455 (14.8%) | 219 (15.1%) | - | - | - | - |

*SMFQ*: Short Mood and Feelings Questionnaire; *CMDs:* Common mental disorders; *CIS-R:* Clinical Interview Schedule – Revised.

*Table S6:* Comparing ALSPAC participants who possess primary care data at each time-point, split by whether they attended/completed each specific data collection event. The numbers with and without ALSPAC data are provided, as are differences in rates of current depression or common mental disorder (CMD) diagnoses from the primary care records. For participants who did not attend the clinic/complete the questionnaire, the age to define a 'current' diagnosis was based on +/- 6 months from the average age each clinic/questionnaire was completed. Individuals who have GP data and completed the clinic/questionnaire, but do not have ALSPAC-derived depression/CMD data (as this session was not completed for whatever reason), are not included in the table below. The number of these individuals at each time point are: 104 at the age 15/16 TF3 clinic (2.8% of those with both ALSPAC and primary care data); 82 at the age 16/17 CCS questionnaire (2.5% of those with both ALSPAC and primary care data); 437 at the age 17/18 TF4 clinic (12.4% of those with both ALSPAC and primary care data); 20 at the age 18/19 CCT questionnaire (1% of those with both ALSPAC and primary care data); 63 at the age 21/22 YPA questionnaire (4.6% of those with both ALSPAC and primary care data); 43 at the age 22/23 YPB questionnaire (3.1% of those with both ALSPAC and primary care data). For a graphical summary of these results, see figure 1.

| Age (time point) | Ave. age at clinic/quest (years (months)) | ALSPAC measure completed | # with and without ALSPAC CMD/ depression data (%) | # with current depression diagnosis in GP records (%) | # with current CMD diagnosis in GP records (%) |
|---|---|---|---|---|---|
| **Age 15/16 (TF3 clinic)** | 15.5 years (186 mths) | Yes | 3,663 (42.8%) | 7 (0.2%) | 26 (0.7%) |
| | | No | 4,902 (57.2%) | 8 (0.2%) | 29 (0.6%) |
| **Age 16/17 (CCS quest)** | 16.67 years (200 mths) | Yes | 3,213 (37.6%) | 17 (0.5%) | 37 (1.2%) |
| | | No | 5,342 (62.4%) | 25 (0.5%) | 43 (0.8%) |
| **Age 17/18 (TF4 clinic)** | 17.83 years (214 mths) | Yes | 3,084 (38.3%) | 30 (1%) | 57 (1.9%) |
| | | No | 4,961 (61.7%) | 45 (0.9%) | 76 (1.5%) |
| **Age 18/19 (CCT quest)** | 18.67 years (224 mths) | Yes | 1,982 (23.6%) | 24 (1.2%) | 53 (2.7%) |
| | | No | 6,402 (76.4%) | 78 (1.2%) | 131 (2.1%) |
| **Age 21/22 (YPA quest)** | 21.92 years (263 mths) | Yes | 1,298 (20.4%) | 35 (2.7%) | 62 (4.8%) |
| | | No | 5,055 (79.6%) | 106 (2.1%) | 155 (3.1%) |
| **Age 22/23 (YPB quest)** | 22/92 years (275 mths) | Yes | 1,325 (23.4%) | 30 (2.3%) | 70 (5.3%) |
| | | no | 4,335 (76.6%) | 81 (1.9%) | 128 (3%) |

*Table S7:* Raw data comparing depression and common mental disorder (CMD) diagnoses based on the Development and Well-Being Assessment (DAWBA) data from the age 15/16 TF3 clinic against various definitions derived from the primary care data at this age (*n*=3,663). Note that values with an asterisk have been suppressed for disclosure control purposes as at least one cell has a value < 5. This table also includes sensitivities, specificities, positive predictive values (PPV) and negative predictive values (NPV) for the depression and CMD diagnoses based on the DAWBA data from this clinic. In these analyses we are treating the ALSPAC data as the reference standard.

| | | **DAWBA Depression** | | | **DAWBA CMD** | | |
|---|---|---|---|---|---|---|---|
| **Primary care definition** | | **No** | **Yes** | **Total** | **No** | **Yes** | **Total** |
| **Current diagnosis** | No | * | * | * | 3,521 | 116 | 3,637 |
| | Yes | * | * | * | 19 | 7 | 26 |
| **Sensitivity** | | * | | | 5.7% (2.3; 11.4) | | |
| **Specificity** | | * | | | 99.5% (99.2; 99.7) | | |
| **Positive Predictive Value** | | * | | | 26.9% (11.6; 47.8) | | |
| **Negative Predictive Value** | | * | | | 96.8% (96.2; 97.4) | | |
| | | **No** | **Yes** | **Total** | **No** | **Yes** | **Total** |
| **Current diagnosis, treated** | No | * | * | * | * | * | * |
| | Yes | * | * | * | * | * | * |
| **Sensitivity** | | * | | | * | | |
| **Specificity** | | * | | | * | | |
| **Positive Predictive Value** | | * | | | * | | |
| **Negative Predictive Value** | | * | | | * | | |
| | | **No** | **Yes** | **Total** | **No** | **Yes** | **Total** |
| **Current diagnosis or symptoms or treatment** | No | 3,576 | 52 | 3,628 | 3,491 | 109 | 3,600 |
| | Yes | 30 | 5 | 35 | 49 | 14 | 63 |
| **Sensitivity** | | 8.8% (2.9; 19.3) | | | 11.4% (6.4; 18.4) | | |
| **Specificity** | | 99.2% (98.8; 99.4) | | | 98.6% (98.2; 99) | | |
| **Positive Predictive Value** | | 14.3% (4.8; 30.3) | | | 22.2% (12.7; 34.5) | | |
| **Negative Predictive Value** | | 98.6% (98.1; 98.9) | | | 97 (96.4; 97.5) | | |

*Table S8:* Raw data comparing depression diagnoses based on the Short Mood and Feelings Questionnaire (SMFQ) data from the age 16/17 CCS questionnaire against various definitions derived from the primary care data at this age (*n*=3,213). Note that values with an asterisk have been suppressed for disclosure control purposes as at least one cell has a value < 5. This table also includes sensitivities, specificities, positive predictive values (PPV) and negative predictive values (NPV) for the depression diagnoses based on the SMFQ data from this questionnaire. In these analyses we are treating the ALSPAC data as the reference standard.

| | | **SMFQ Depression** | | |
|---|---|---|---|---|
| **Primary care definition** | | **No** | **Yes** | **Total** |
| **Current diagnosis** | No | 2,719 | 477 | 3,196 |
| | Yes | 9 | 8 | 17 |
| **Sensitivity** | | 1.6% (0.7; 3.2) | | |
| **Specificity** | | 99.7% (99.4; 99.8) | | |
| **Positive Predictive Value** | | 47.1% (23; 72.2) | | |
| **Negative Predictive Value** | | 85.1% (83.8; 86.3) | | |
| | | **No** | **Yes** | **Total** |
| **Current diagnosis, treated** | No | * | * | * |
| | Yes | * | * | * |
| **Sensitivity** | | * | | |
| **Specificity** | | * | | |
| **Positive Predictive Value** | | * | | |
| **Negative Predictive Value** | | * | | |
| | | **No** | **Yes** | **Total** |
| **Current diagnosis or symptoms or treatment** | No | 2,694 | 451 | 3,145 |
| | Yes | 34 | 34 | 68 |
| **Sensitivity** | | 7% (4.9; 9.7) | | |
| **Specificity** | | 98.8% (98.3; 99.1) | | |
| **Positive Predictive Value** | | 50% (37.6; 62.4) | | |
| **Negative Predictive Value** | | 85.7% (84.4; 86.9) | | |

*Table S9:* Raw data comparing depression diagnoses based on the Short Mood and Feelings Questionnaire (SMFQ) data from the age 18/19 CCT questionnaire against various definitions derived from the primary care data at this age (*n*=1,982). This table also includes sensitivities, specificities, positive predictive values (PPV) and negative predictive values (NPV) for the depression diagnoses based on the SMFQ data from this questionnaire. In these analyses we are treating the ALSPAC data as the reference standard.

| | | **SMFQ Depression** | | |
|---|---|---|---|---|
| **Primary care definition** | | **No** | **Yes** | **Total** |
| **Current diagnosis** | No | 1,586 | 372 | 1,958 |
| | Yes | 9 | 15 | 24 |
| **Sensitivity** | | 3.9% (2.2; 6.3) | | |
| **Specificity** | | 99.4% (98.9; 99.7) | | |
| **Positive Predictive Value** | | 62.5% (40.6; 81.2) | | |
| **Negative Predictive Value** | | 81% (79.2; 82.7) | | |
| | | **No** | **Yes** | **Total** |
| **Current diagnosis, treated** | No | 1,590 | 374 | 1,964 |
| | Yes | 5 | 13 | 18 |
| **Sensitivity** | | 3.4% (1.8; 5.7) | | |
| **Specificity** | | 99.7% (99.3; 99.9) | | |
| **Positive Predictive Value** | | 72.2% (46.5; 90.3) | | |
| **Negative Predictive Value** | | 81% (79.1; 82.7) | | |
| | | **No** | **Yes** | **Total** |
| **Current diagnosis or symptoms or treatment** | No | 1,542 | 320 | 1,862 |
| | Yes | 53 | 67 | 120 |
| **Sensitivity** | | 17.3% (13.7; 21.5) | | |
| **Specificity** | | 96.7% (95.7; 97.5) | | |
| **Positive Predictive Value** | | 55.8% (46.5; 64.9) | | |
| **Negative Predictive Value** | | 82.8% (81; 84.5) | | |

*Table S10:* Raw data comparing depression diagnoses based on the Short Mood and Feelings Questionnaire (SMFQ) data from the age 21/22 YPA questionnaire against various definitions derived from the primary care data at this age (*n*=1,298). This table also includes sensitivities, specificities, positive predictive values (PPV) and negative predictive values (NPV) for the depression diagnoses based on the SMFQ data from this questionnaire. In these analyses we are treating the ALSPAC data as the reference standard.

| | | **SMFQ Depression** | | |
|---|---|---|---|---|
| **Primary care definition** | | **No** | **Yes** | **Total** |
| **Current diagnosis** | No | 1,082 | 181 | 1,263 |
| | Yes | 16 | 19 | 35 |
| **Sensitivity** | | 9.5% (5.8; 14.4) | | |
| **Specificity** | | 98.5% (97.6; 99.2) | | |
| **Positive Predictive Value** | | 54.3% (36.6; 71.2) | | |
| **Negative Predictive Value** | | 85.7% (83.6; 87.6) | | |
| | | **No** | **Yes** | **Total** |
| **Current diagnosis, treated** | No | 1,086 | 182 | 1,268 |
| | Yes | 12 | 18 | 30 |
| **Sensitivity** | | 9% (5.4; 13.9) | | |
| **Specificity** | | 98.9% (98.1; 99.4) | | |
| **Positive Predictive Value** | | 60% (40.6; 77.3) | | |
| **Negative Predictive Value** | | 85.6% (83.6; 87.5) | | |
| | | **No** | **Yes** | **Total** |
| **Current diagnosis or symptoms or treatment** | No | 1,026 | 132 | 1,158 |
| | Yes | 72 | 68 | 140 |
| **Sensitivity** | | 34% (27.5; 41) | | |
| **Specificity** | | 93.4% (91.8; 94.8) | | |
| **Positive Predictive Value** | | 48.6% (40; 57.2) | | |
| **Negative Predictive Value** | | 88.6% (86.6; 90.4) | | |

*Table S11:* Raw data comparing depression diagnoses based on the Short Mood and Feelings Questionnaire (SMFQ) data from the age 22/23 YPB questionnaire against various definitions derived from the primary care data at this age (*n*=1,326). This table also includes sensitivities, specificities, positive predictive values (PPV) and negative predictive values (NPV) for the depression diagnoses based on the SMFQ data from this questionnaire. In these analyses we are treating the ALSPAC data as the reference standard.

| | | **SMFQ Depression** | | |
|---|---|---|---|---|
| **Primary care definition** | | **No** | **Yes** | **Total** |
| **Current diagnosis** | No | 1,093 | 202 | 1,295 |
| | Yes | 10 | 20 | 30 |
| **Sensitivity** | | 9% (5.6; 13.6) | | |
| **Specificity** | | 99.1% (98.3; 99.6) | | |
| **Positive Predictive Value** | | 66.7% (47.2; 82.7) | | |
| **Negative Predictive Value** | | 84.4% (82.3; 86.3) | | |
| | | **No** | **Yes** | **Total** |
| **Current diagnosis, treated** | No | 1,095 | 205 | 1,300 |
| | Yes | 8 | 17 | 25 |
| **Sensitivity** | | 7.7% (4.5; 12) | | |
| **Specificity** | | 99.3% (98.6; 99.7) | | |
| **Positive Predictive Value** | | 68% (46.5; 85.1) | | |
| **Negative Predictive Value** | | 84.2% (82.1; 86.2) | | |
| | | **No** | **Yes** | **Total** |
| **Current diagnosis or symptoms or treatment** | No | 1,035 | 161 | 1,196 |
| | Yes | 68 | 61 | 129 |
| **Sensitivity** | | 27.5% (21.7; 33.9) | | |
| **Specificity** | | 93.8% (92.2; 95.2) | | |
| **Positive Predictive Value** | | 47.3% (38.4; 56.3) | | |
| **Negative Predictive Value** | | 86.5% (84.5; 88.4) | | |

*Table S12:* Results of the identification in primary care records analysis, based on whether individuals who were diagnosed have having depression or common mental disorders (CMDs) in ALSPAC were also diagnosed based on primary care record data (with primary care diagnosis defined as 'current diagnosis or treatment or symptoms'). Odds ratios are displayed (with 95% confidence intervals). Due to the small sample sizes in some analyses these estimates are rather imprecise, especially regarding the age 15/16 TF3 clinic as very few individuals were classified correctly/diagnosed as depressed/CMD in primary care records. Coefficients are odds ratios derived from univariable logistic regressions and denote the odds of identification relative to the baseline (e.g., for age 17/18 TF4 clinic depression, females have three times greater odds of being identified than males). Note also that when comparing against ALSPAC data (e.g., mother's marital status, parental education, etc.) the sample size for each analysis will vary as the variables come from different data sources, with different levels of completeness. For a graphical illustration of key results, see figure S1.

| *Variable* | *Age 15/16 TF3 clinic* | | *Age 16/17 CCS questionnaire* | *Age 17/18 TF4 clinic* | | *Age 18/19 CCT questionnaire* | *Age 21/22 YPA questionnaire* | *Age 22/23 YPB questionnaire* |
|---|---|---|---|---|---|---|---|---|
| | *Dep. (n = 57)* | *CMD (n = 123)* | *Dep. (n = 485)* | *Dep. (n = 243)* | *CMD (n = 455)* | *Dep. (n = 387)* | *Dep. (n = 200)* | *Dep. (n = 222)* |
| *Sex (ref = Male)* | 0.55 (0.08; 3.66) | 0.71 (0.2; 2.45) | 1.38 (0.56; 3.43) | 3.30 (1.33; 8.14) | 3.14 (1.61; 6.13) | 0.83 (0.44; 1.55) | 1.54 (0.78; 3.04) | 1.34 (0.68; 2.63) |
| *Age at data collection event (months)* | 0.81 (0.55; 1.19) | 0.89 (0.73; 1.08) | 0.93 (0.81; 1.08) | 0.98 (0.92; 1.06) | 0.98 (0.92; 1.03) | 0.98 (0.94; 1.03) | 1.02 (0.97; 1.07) | 0.98 (0.94; 1.03) |
| *Mother's age at birth (years)* | 0.89 (0.72; 1.11) | 0.91 (0.8; 1.04) | 0.96 (0.89; 1.04) | 0.95 (0.89; 1.01) | 0.96 (0.91; 1.01) | 1.01 (0.96; 1.07) | 0.9 (0.84; 0.97) | 1 (0.93; 1.06) |
| *Mother's home ownership status (ref = owned/mort.)* | 0.63 (0.06; 6.09) | 2.23 (0.65; 7.68) | 0.91 (0.34; 2.44) | 1.65 (0.81; 3.35) | 1.31 (0.73; 2.34) | 1.41 (0.71; 2.8) | 0.92 (0.43; 1.99) | 0.8 (0.32; 1.98) |
| *Mother's marital status (ref = married)* | 1.38 (0.21; 9.07) | 2.61 (0.8; 8.48) | 0.77 (0.29; 2.06) | 1.48 (0.76; 2.87) | 0.98 (0.57; 1.72) | 1.25 (0.67; 2.36) | 1.43 (0.68; 3.01) | 1.02 (0.45; 2.28) |
| *Mother's age at first pregnancy (years)* | 1.16 (0.88; 1.53) | 0.89 (0.77; 1.03) | 1 (0.93; 1.08) | 0.93 (0.87; 0.99) | 0.96 (0.91; 1) | 1 (0.95; 1.05) | 0.97 (0.91; 1.02) | 0.99 (0.93; 1.05) |
| *Mother's parity at child's birth (ref = first child)* | 0.38 (0.04; 3.91) | 1.49 (0.41; 5.4) | 0.68 (0.32; 1.43) | 0.92 (0.5; 1.7) | 1.2 (0.73; 1.96) | 1.26 (0.72; 2.21) | 0.84 (0.45; 1.56) | 1.04 (0.56; 1.92) |
| *Mother's highest education level (ref = CSE/Voc./O level)* | NC | 0.35 (0.07; 1.66) | 0.83 (0.38; 1.81) | 0.78 (0.41; 1.47) | 1.09 (0.66; 1.8) | 0.9 (0.51; 1.57) | 1.1 (0.58; 2.11) | 1.26 (0.66; 2.4) |
| *Father's highest education level (ref = CSE/Voc./O level)* | 4.06 (0.39; 42.49) | 2.32 (0.63; 8.51) | 0.77 (0.35; 1.7) | 1.16 (0.61; 2.21) | 1.18 (0.71; 1.98) | 1.2 (0.68; 2.14) | 0.85 (0.45; 1.61) | 0.67 (0.36; 1.26) |
| *Mother's social class (ref = I/II)* | NC | NC | 0.96 (0.44; 2.1) | 1.33 (0.67; 2.65) | 0.86 (0.5; 1.47) | 0.9 (0.5; 1.64) | 0.77 (0.39; 1.54) | 0.43 (0.22; 0.85) |
| *Father's social class (ref = I/II)* | 0.56 (0.08; 3.66) | 1.04 (0.33; 3.35) | 1.74 (0.76; 3.99) | 1.57 (0.81; 3.05) | 1 (0.59; 1.68) | 1.24 (0.7; 2.21) | 1.31 (0.69; 2.48) | 0.77 (0.4; 1.47) |
| *Child's ethnic background (ref = white)* | NC | 0.91 (0.1; 7.9) | 0.87 (0.11; 6.8) | 0.37 (0.04; 2.99) | 0.67 (0.19; 2.32) | 0.75 (0.16; 3.42) | 1.23 (0.2; 7.59) | 1.22 (0.3; 5.06) |
| *Child's education (# A*-C GCSE equivalents)* | 0.93 (0.72; 1.22) | 0.92 (0.79; 1.06) | 0.89 (0.8; 0.98) | 0.97 (0.9; 1.05) | 0.99 (0.92; 1.05) | 0.94 (0.88; 1.01) | 1.02 (0.94; 1.1) | 1 (0.92; 1.08) |
| *CIS-R depression score (ref = 1 for depression model; ref = 0 for CMD model)* | NA | NA | NA | 2: 1.44 (0.54; 3.83)<br>3: 2.21 (0.88; 5.54)<br>4: 3.78 (1.5; 9.53) | 1: 1.97 (0.83; 4.7)<br>2: 2.85 (1.21; 6.67)<br>3: 5.73 (2.57; 12.8) | NA | NA | NA |

| | | | | | | | | |
|---|---|---|---|---|---|---|---|---|
| | | | | | 4: 10.08 (4.4; 23.09) | | | |
| *CIS-R diagnosis of moderate depressive episode (ref = no)* | NA | NA | NA | 2.49 (1.26; 4.92) | 3.23 (2; 5.22) | NA | NA | NA |
| *CIS-R diagnosis of severe depressive episode (ref = no)* | NA | NA | NA | 2.96 (1.39; 6.29) | 5.01 (2.44; 10.3) | NA | NA | NA |
| *CIS-R GAD symptoms (ref = no)* | NA | NA | NA | 2.11 (1.15; 3.88) | 1.49 (0.93; 2.38) | NA | NA | NA |
| *CIS-R total score* | NA | NA | NA | 1.09 (1.04; 1.15) | 1.11 (1.07; 1.15) | NA | NA | NA |
| *DAWBA predicted depression (ref = ~50%)* | >70%: 1.38 (0.14; 13.95) | ~0.5%: 0.29 (0.03; 2.58)<br>~15%: 2.32 (0.67; 8.05)<br>>70%: 1.08 (0.11; 10.47) | NA | NA | NA | NA | NA | NA |
| *DAWBA predicted anxiety (ref = ~50%)* | ~3%: 2.53 (0.25; 25.39) | ~3%: 1.45 (0.41; 5.15) | NA | NA | NA | NA | NA | NA |
| *Total SMFQ score* | NA | NA | 1.11 (1.02; 1.21) | NA | NA | 1.09 (1.03; 1.16) | 1.14 (1.06; 1.22) | 1.08 (1.01; 1.16) |
| *Historical depression diagnosis* | NC | NC | 14.03 (1.91; 102.9) | 8.12 (2.03; 32.52) | 14.56 (4.57; 46.38) | 13.86 (4.2; 45.71) | 3.27 (1.52; 7.03) | 3.07 (1.46; 6.46) |
| *Historical anxiety diagnosis* | 34 (2.36; 489.96) | 8.92 (1.15; 69.18) | 7.22 (2.55; 20.43) | 2.34 (0.71; 7.68) | 2.24 (0.97; 5.18) | 6.09 (2.47; 15.01) | 2.81 (1.31; 6.01) | 1.95 (0.87; 4.33) |
| *Current anxiety diagnosis* | NC | NA | 13.59 (4.28; 43.16) | 16.94 (1.94; 148.16) | NA | 9.76 (3.68; 25.87) | 10.16 (2.79; 37.09) | 5.07 (1.75; 14.63) |
| *Historical depression symptoms* | NC | NC | 4.69 (1.6; 13.7) | 5.5 (2.21; 13.65) | 5.34 (2.58; 11.08) | 4.71 (2.29; 9.68) | 5.6 (2.86; 10.97) | 4.75 (2.52; 8.97) |
| *Historical somatic symptoms* | 4.08 (0.34; 48.86) | 8.32 (1.92; 36.05) | 3.77 (1.51; 9.4) | 1.72 (0.82; 3.61) | 1.66 (0.92; 2.98) | 2.53 (1.38; 4.66) | 3.77 (2.01; 7.07) | 2.59 (1.37; 4.9) |
| *Current somatic symptoms* | NC | 1.32 (0.15; 11.85) | 2.47 (0.69; 8.89) | 3.5 (1.32; 9.29) | 6.18 (2.78; 13.76) | 3.73 (1.65; 8.46) | 1.78 (0.62; 5.13) | 12.9 (3.5; 47.57) |
| *Historical general symptoms* | 0.68 (0.07; 6.6) | 1.78 (0.51; 6.26) | 2.63 (1.27; 5.45) | 3.35 (1.79; 6.27) | 3.46 (2.07; 5.75) | 4.04 (2.32; 7.06) | 3.62 (1.91; 6.88) | 1.79 (0.97; 3.31) |
| *Current general symptoms* | 3 (0.27; 33.64) | 1.12 (0.13; 9.85) | 2.2 (0.62; 7.84) | 3.99 (1.29; 12.4) | 2.38 (1.02; 5.54) | 2.15 (0.9; 5.14) | 3.67 (1.04; 13.02) | 3.35 (1.08; 10.4) |
| *Historical anti-depressant treatment* | NC | NC | 11.89 (3.04; 46.61) | 3.97 (1.53; 10.31) | 7.46 (3.22; 17.26) | 23.37 (9.47; 57.66) | 9.87 (5; 19.44) | 9.59 (4.83; 19.04) |
| *Historical anti-anxiety treatment* | NC | 8.31 (0.49; 140.9) | 3.46 (0.71; 26.98) | 1.8 (0.58; 5.6) | 3.74 (1.5; 9.34) | 6.24 (2.78; 14.01) | 4.03 (1.9; 8.55) | 5.49 (2.68; 11.27) |
| *Current anti-anxiety treatment* | NC | NA | 6.98 (1.23; 39.59) | 6.8 (2.39; 19.32) | NA | 12.9 (5.02; 33.15) | 10.16 (2.79; 37.08) | 5.4 (1.73; 16.84) |
| *# of GP consultations per year* | 1.07 (0.86; 1.34) | 1.06 (0.93; 1.2) | 1.17 (1.11; 1.23) | 1.25 (1.16; 1.34) | 1.24 (1.17; 1.31) | 1.13 (1.07; 1.18) | 1.25 (1.15; 1.35) | 1.23 (1.15; 1.32) |
| *# of GP prescriptions per year* | 0.93 (0.64; 1.37) | 0.88 (0.69; 1.11) | 1.22 (1.11; 1.35) | 1.25 (1.14; 1.38) | 1.29 (1.19; 1.39) | 1.25 (1.14; 1.36) | 1.44 (1.26; 1.64) | 1.37 (1.22; 1.54) |
| *Asthma diagnosis* | 1.64 (0.25; 10.85) | 1.28 (0.37; 4.41) | 1.77 (0.83; 3.76) | 0.74 (0.37; 1.48) | 0.77 (0.44; 1.36) | 1.44 (0.79; 2.61) | 1 (0.51; 1.94) | 1.82 (0.95; 3.51) |
| *Eczema diagnosis* | 1.07 (0.16; 6.95) | 0.57 (0.17; 1.93) | 1.6 (0.79; 3.27) | 1.35 (0.71; 2.55) | 1.13 (0.69; 1.85) | 1.36 (0.79; 2.34) | 1.18 (0.65; 2.14) | 0.72 (0.39; 1.33) |
| *Eating disorder* | NC | 5.89 (0.89; 38.83) | 5.58 (1.04; 29.87) | 3.31 (0.92; 11.88) | 5.28 (1.73; 16.13) | 2.27 (0.76; 6.75) | 1.12 (0.32; 3.95) | 1.96 (0.6; 6.44) |

| | | | | | | | | |
|---|---|---|---|---|---|---|---|---|
| *History of mental health issues* | NC | 2.72 (0.26; 28.08) | 1.49 (0.18; 12.11) | 12.25 (2.47; 60.76) | 7.96 (2.28; 27.85) | 8.54 (2.7; 27.02) | 8.67 (1.79; 42.05) | 6.83 (1.71; 27.34) |
| *Self-harm* | 5.11 (0.7; 37.06) | 3.97 (0.9; 17.61) | 4.15 (1.44; 11.97) | 3.72 (1.61; 8.57) | 4.46 (2.21; 9.01) | 3.61 (1.7; 7.69) | 3.23 (1.3; 8) | 4.31 (1.56; 11.92) |
| *Any other mental health issues* | NC | 9.64 (1.73; 53.63) | 2.47 (0.69; 8.89) | 1.78 (0.63; 5.04) | 2.86 (1.25; 6.54) | 2.08 (0.83; 5.23) | 1.12 (0.45; 2.83) | 1.58 (0.63; 3.99) |
| *Referral to mental health services* | 1.92 (0.18; 20.11) | 7.01 (1.89; 25.97) | 3.39 (1.37; 8.4) | 5.84 (2.83; 12.07) | 5.76 (3.19; 10.42) | 6.85 (3.45; 13.61) | 3.71 (1.79; 7.65) | 4.22 (2.08; 8.56) |
| *Smoking (or ex-smoker)* | NC | 2.1 (0.4; 11.08) | 2.08 (0.9; 4.82) | 2.92 (1.54; 5.52) | 3.38 (2.01; 5.71) | 2.26 (1.26; 4.05) | 1.24 (0.67; 2.28) | 1.15 (0.61; 2.16) |

NA = Not applicable (either because measure not available at that time-point, or because the variable is part of the GP diagnosis variable; e.g., current anti-anxiety treatment is part of the 'current diagnosis or symptoms or treatment' definition of CMDs)

NC = Not calculable (due to collinearity in model as a result of lack of data). Variables where half or more of all analyses were not calculable due to collinearity have been removed from this table, as sample sizes are likely to be too low to be informative. These variables are: historical phobia diagnosis, current phobia diagnosis, historical anxiety symptoms, current anxiety symptoms, ADHD, conduct disorder, autism spectrum disorder, substance abuse, family history of mental illness, family history of depression, personal history of depression and other psychological illness.

*Figure S1 (following page):* Graphical summary of key results of the identification in primary care records analysis, based on whether individuals who were diagnosed have having depression or common mental disorders (CMDs) in ALSPAC were also diagnosed based on primary care record data (with primary care diagnosis defined as 'current diagnosis or treatment or symptoms'). Values for depression are displayed in black, and common mental disorders are in red. Odds ratios and 95% confidence intervals are displayed on the y-axis (on the log scale), with time point along the x-axis (going forwards in time, from the age 15/16 TF3 clinic to the age 22/23 YPB questionnaire. Due to the small sample sizes in some analyses these estimates are rather imprecise, especially regarding the 15/16 clinic as very few individuals were classified correctly/diagnosed as depressed/CMD in primary care records. Coefficients are odds ratios derived from univariable logistic regressions and denote the odds of identification relative to the baseline (e.g., for age 17/18 TF4 clinic depression, females have three times greater odds of being identified than males). Note also that when comparing against ALSPAC data (e.g., mother's marital status, parental education, etc.) the sample size for each analysis will vary as the variables come from different data sources, with different levels of completeness. For a full list of results, see table S11.

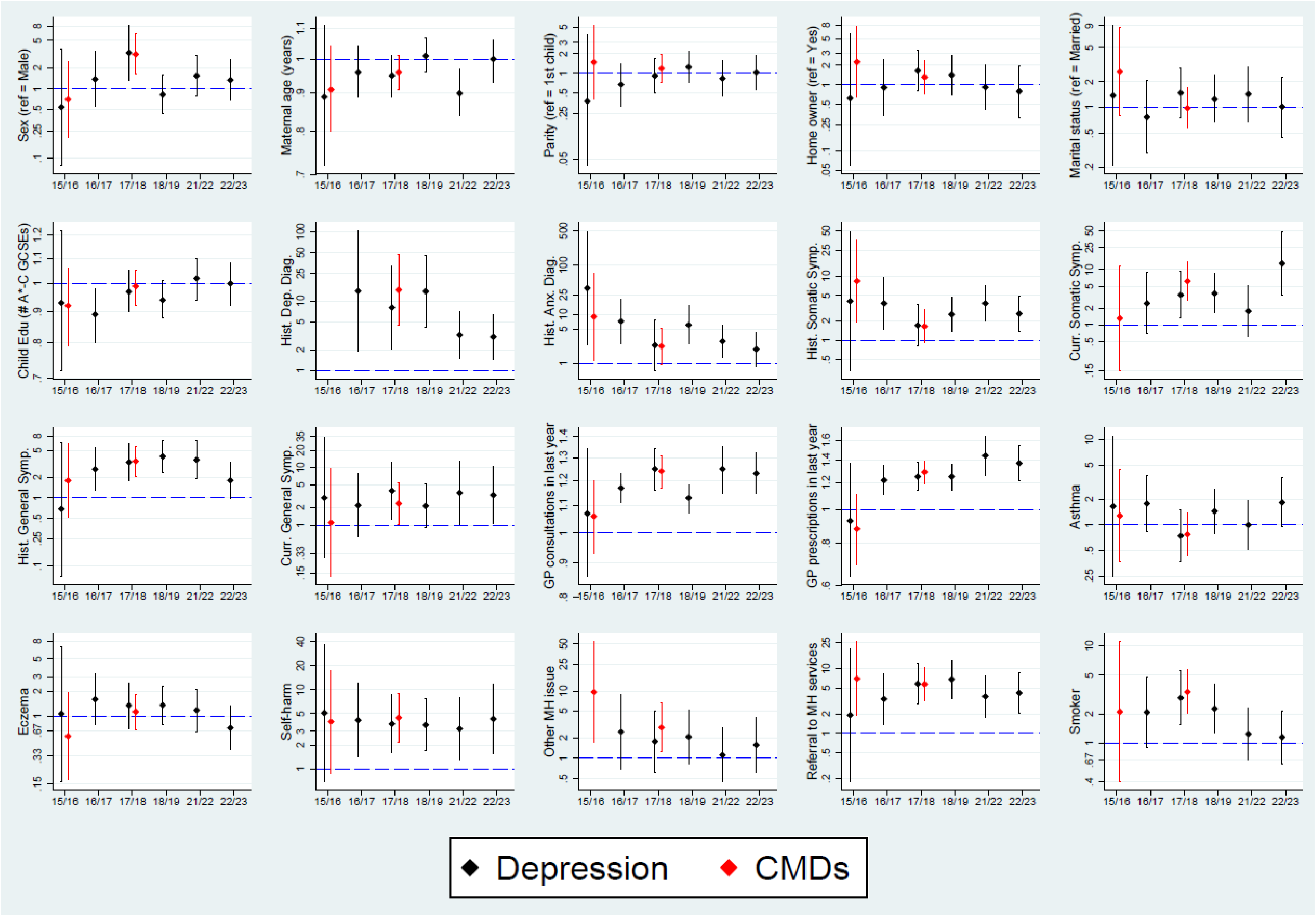
Sex (ref = Male)
Maternal age (years)
Parity (ref = 1st child)
Home owner (ref = Yes)
Marital status (ref = Married)
Child Edu (# A*-C GCSEs)
Hist. Dep. Diag.
Hist. Anx. Diag.
Hist. Somatic Symp.
Curr. Somatic Symp.
Hist. General Symp.
Curr. General Symp.
GP consultations in last year
GP prescriptions in last year
Asthma
Eczema
Self-harm
Other MH issue
Referral to MH services
Smoker
15/16
16/17
17/18
18/19
21/22
22/23
Depression
CMDs

*Table S13:* Penalised coefficients from the optimal lasso prediction models, predicting ALSPAC questionnaire-derived depression or common mental disorder (CMD) cases using only linked primary care record data. Coefficients are log-odds estimates. Note that as the estimates are derived from lasso models used for model prediction, standard errors are not calculated.

| | **Age 15/16 TF3 clinic** | | **Age 16/17 CCS quest** | **Age 17/18 TF4 clinic** | | **Age 18/19 CCT quest** | **Age 21/22 YPA quest** | **Age 22/23 YPB quest** |
|---|---|---|---|---|---|---|---|---|
| | Dep. | CMD | Dep. | Dep. | CMD | Dep. | Dep. | Dep. |
| **Constant** | -4.307 | -3.788 | -2.602 | -3.24 | -5.966 | -1.864 | -2.036 | -5.805 |
| **Self-harm status (ref: no)** | 2.297 | 1.225 | 0.476 | 1.469 | 1.301 | 1.536 | - | 0.538 |
| **Historical general symptoms (ref: no)** | 0.464 | 0.143 | 0.204 | - | - | - | - | - |
| **Mean number of GP consultations** | 0.02 | 0.036 | 0.03 | 0.046 | 0.019 | 0.004 | - | - |
| **Historical depression diagnosis (ref: no)** | 0.84 | - | - | - | - | - | 0.421 | 0.071 |
| **Current depression diagnosis (ref: no)** | 0.478 | 1.445 | 0.217 | 0.472 | 0.68 | 0.69 | - | 0.606 |
| **Sex (ref: male)** | - | 0.544 | 0.777 | 0.437 | 0.493 | 0.449 | - | - |
| **Eating disorder (ref: no)** | - | 0.748 | - | 0.791 | - | - | - | - |
| **Referral to mental health services (ref: no)** | - | 0.242 | 1.042 | - | 0.525 | 0.22 | 0.539 | 0.29 |
| **Current phobia diagnosis (ref: no)** | - | 1.434 | - | - | - | 2.281 | - | - |
| **Current anxiety diagnosis (ref: no)** | - | 0.349 | 1.035 | - | 0.016 | 1.057 | - | - |
| **Current antidepressant use (ref: no)** | - | 0.312 | - | 0.808 | 0.618 | 0.725 | 0.73 | 0.82 |
| **Current depression symptoms (ref: no)** | - | - | 0.218 | 1.156 | 0.76 | - | 0.033 | 0.502 |
| **Smoking status (ref: no)** | - | - | 0.466 | 0.058 | - | - | - | - |
| **Historical depression symptoms (ref: no)** | - | - | 0.469 | - | 0.388 | 0.457 | 0.18 | 0.54 |
| **Historical somatic symptoms (ref: no)** | - | - | 0.125 | - | 0.185 | 0.016 | - | - |
| **Autism spectrum disorder (ref: no)** | - | - | 2.131 | - | - | - | - | 0.702 |
| **Family history of depression (ref: no)** | - | - | 2.61 | - | - | - | - | - |
| **Conduct disorder (ref: no)** | - | - | 0.89 | - | - | - | - | - |
| **ADHD (ref: no)** | - | - | -0.622 | - | - | - | - | - |
| **Substance abuse (ref: no)** | - | - | 0.881 | - | - | - | - | - |
| **Historical phobia diagnosis (ref: no)** | - | - | 0.373 | - | - | - | - | 0.447 |
| **Historical anxiety symptoms (ref: no)** | - | - | 0.292 | - | - | - | - | -0.634 |
| **Historical anti-anxiety meds (ref: no)** | - | - | - | 0.63 | 0.17 | 0.426 | - | - |
| **Current anti-anxiety meds (ref: no)** | - | - | - | 0.251 | - | - | 0.122 | - |
| **Eczema (ref: no)** | - | - | - | 0.027 | - | - | - | - |

| | | | | | | | | |
|---|---|---|---|---|---|---|---|---|
| **Historical antidepressant use (ref: no)** | - | - | - | 0.058 | 0.135 | -0.066 | 0.613 | 0.688 |
| **Historical anxiety diagnosis (ref: no)** | - | - | - | - | 0.524 | - | - | - |
| **Age (months)** | - | - | - | - | 0.017 | - | - | 0.014 |
| **Current general symptoms (ref: no)** | - | - | - | - | 0.189 | - | - | - |
| **Current somatic symptoms (ref: no)** | - | - | - | - | - | 0.613 | - | - |
| **History of depression (ref: no)** | - | - | - | - | - | 0.808 | - | - |
| **History of mental health issues (ref: no)** | - | - | - | - | - | -0.198 | - | - |
| **Other psychological illness (ref: no)** | - | - | - | - | - | -0.013 | - | - |

*Table S14:* Full models to estimate the predicted probability of ALSPAC questionnaire-derived depression or common mental disorders (CMDs) at each time-point, based on the best-fitting lasso model using only linked primary care record data as predictors. Code is provided in Stata syntax.

| **Time-point (*n*)** | **Diagnosis** | **Model to estimate predicted probability of diagnosis** |
|---|---|---|
| **Age 15/16 TF3 clinic (*n* = 3,663)** | Depression | gen TF3_dep_DAWBA = (exp(-4.306898 + (selfharm_TF3 * 2.297141) + (hist_gensym_TF3 * 0.4635765) + (consult_mean_TF3 * 0.0201705) + (hist_depress_TF3 * 0.8404444) + (curr_depress_TF3 * 0.4780882))) / (1 (exp(-4.306898 + (selfharm_TF3 * 2.297141) + (hist_gensym_TF3 * 0.4635765) + (consult_mean_TF3 * 0.0201705) + (hist_depress_TF3 * 0.8404444) + (curr_depress_TF3 * 0.4780882)))) |
| | CMD | gen TF3_CMD_DAWBA = (exp(-3.78843 + (sex * 0.5437601) + (selfharm_TF3 * 1.224612) + (consult_mean_TF3 * 0.0356085) + (curr_depress_TF3 * 1.444507) + (eatdisorder_TF3 * 0.7475241) + (hist_gensym_TF3 * 0.1431262) + (referral_TF3 * 0.2418507) + (curr_phob_TF3 * 1.434107) + (curr_anx_TF3 * 0.3485559) + (curr_depdrugs_TF3 * 0.3121066))) / (1 + (exp(-3.78843 + (sex * 0.5437601) + (selfharm_TF3 * 1.224612) + (consult_mean_TF3 * 0.0356085) + (curr_depress_TF3 * 1.444507) + (eatdisorder_TF3 * 0.7475241) + (hist_gensym_TF3 * 0.1431262) + (referral_TF3 * 0.2418507) + (curr_phob_TF3 * 1.434107) + (curr_anx_TF3 * 0.3485559) + (curr_depdrugs_TF3 * 0.3121066)))) |
| **Age 16/17 CCS quest (*n* = 3,213)** | Depression | gen CCS_dep_SMFQ = (exp(-2.602106 + (sex * 0.7768598) + (referral_CCS * 1.042151) + (asd_CCS * 2.131247) + (smoke_CCS * 0.4664435) + (consult_mean_CCS * 0.0301841) + (curr_anx_CCS * 1.03486) + (fhdep_CCS * 2.610094) + (hist_gensym_CCS * 0.203526) + (selfharm_CCS * 0.4762654) + (conduct_CCS * 0.8898849) + (hist_depsymp_CCS * 0.4685372) + (adhd_CCS * -0.6223789) + (hist_somsym_CCS * 0.1245751) + (alcdrug_CCS * 0.8809454) + (hist_phob_CCS * 0.3725191) + (curr_depsymp_CCS * 0.2181478) + (curr_depress_CCS * 0.2167674) + (hist_anxsymp_CCS * 0.2916632))) / (1 + (exp(-2.602106 + (sex * 0.7768598) + (referral_CCS * 1.042151) + (asd_CCS * 2.131247) + (smoke_CCS * 0.4664435) + (consult_mean_CCS * 0.0301841) + (curr_anx_CCS * 1.03486) + (fhdep_CCS * 2.610094) + (hist_gensym_CCS * 0.203526) + (selfharm_CCS * 0.4762654) + (conduct_CCS * 0.8898849) + (hist_depsymp_CCS * 0.4685372) + (adhd_CCS * -0.6223789) + (hist_somsym_CCS * 0.1245751) + (alcdrug_CCS * 0.8809454) + (hist_phob_CCS * 0.3725191) + (curr_depsymp_CCS * 0.2181478) + (curr_depress_CCS * 0.2167674) + (hist_anxsymp_CCS * 0.2916632))) ) |
| **Age 17/18 TF4 clinic (*n* = 3,084)** | Depression | gen TF4_dep_CISR = (exp(-3.239794 + (sex * 0.436772) + (selfharm_TF4 * 1.469416) + (consult_mean_TF4 * 0.0459718) + (referral_TF4 * 0.7906288) + (curr_depsymp_TF4 * 1.155598) + (curr_depdrugs_TF4 * 0.8078427) + (hist_anxdrugs_TF4 * 0.6298965) + (curr_depress_TF4 * 0.4717727) + (curr_anxdrugs_TF4 * 0.2511227) + (smoke_TF4 * 0.0582065) + (eczema * 0.0274914) + (hist_depdrugs_TF4 * 0.0581372))) / (1 + (exp(-3.239794 + (sex * 0.436772) + (selfharm_TF4 * 1.469416) + (consult_mean_TF4 * 0.0459718) + (referral_TF4 * 0.7906288) + (curr_depsymp_TF4 * 1.155598) + (curr_depdrugs_TF4 * 0.8078427) + (hist_anxdrugs_TF4 * 0.6298965) + (curr_depress_TF4 * 0.4717727) + (curr_anxdrugs_TF4 * 0.2511227) + (smoke_TF4 * 0.0582065) + (eczema * 0.0274914) + (hist_depdrugs_TF4 * 0.0581372))) ) |
| | CMD | gen TF4_CMD_CISR = (exp(-5.96604 + (sex * 0.4931608) + (selfharm_TF4 * 1.301296) + (referral_TF4 * 0.5245662) + (curr_depsymp_TF4 * 0.7602904) + (curr_depdrugs_TF4 * 0.6179576) + (consult_mean_TF4 * 0.0194573) + (hist_anx_TF4 * |

| | | |
|---|---|---|
|  |  | 0.5237928) + (TF4_agemths * 0.0168863) + + (curr_depress_TF4 * 0.6795285) + (hist_depsymp_TF4 * 0.3876382) + (hist_somsym_TF4 * 0.184908) + (curr_gensym_TF4 * 0.189229) + (hist_anxdrugs_TF4 * 0.1699545) + (hist_depdrugs_TF4 * 0.1345062) + (curr_anx_TF4 * 0.0156222))) / (1 + (exp(-5.96604 + (sex * 0.4931608) + (selfharm_TF4 * 1.301296) + (referral_TF4 * 0.5245662) + (curr_depsymp_TF4 * 0.7602904) + (curr_depdrugs_TF4 * 0.6179576) + (consult_mean_TF4 * 0.0194573) + (hist_anx_TF4 * 0.5237928) + (TF4_agemths * 0.0168863) + + (curr_depress_TF4 * 0.6795285) + (hist_depsymp_TF4 * 0.3876382) + (hist_somsym_TF4 * 0.184908) + (curr_gensym_TF4 * 0.189229) + (hist_anxdrugs_TF4 * 0.1699545) + (hist_depdrugs_TF4 * 0.1345062) + (curr_anx_TF4 * 0.0156222))) ) |
| **Age 18/19 CCT quest (*n* = 1,982)** | Depression | gen CCS_dep_SMFQ = (exp(-1.864214 + (selfharm_CCT * 1.536287) + (sex * 0.4488152) + (curr_depdrugs_CCT * 0.7249568) + (curr_anx_CCT * 1.056577) + (curr_somsym_CCT * 0.6129295) + (curr_phob_CCT * 2.280971) + (hist_depsymp_CCT * 0.4572833) + (hist_anxdrugs_CCT * 0.4263218) + (curr_depress_CCT * 0.6901121) + (referral_CCT * 0.220459) + (histdep_CCT * 0.807517) + (histmh_CCT * -0.197546) + (consult_mean_CCT * 0.0.0041373) + (hist_depdrugs_CCT * -0.0660863) + (hist_somsym_CCT * 0.015973) + (CCT_agemths * -0.0003042) + (psych_other_CCT * -0.0130987))) / (1 + (exp(-1.864214 + (selfharm_CCT * 1.536287) + (sex * 0.4488152) + (curr_depdrugs_CCT * 0.7249568) + (curr_anx_CCT * 1.056577) + (curr_somsym_CCT * 0.6129295) + (curr_phob_CCT * 2.280971) + (hist_depsymp_CCT * 0.4572833) + (hist_anxdrugs_CCT * 0.4263218) + (curr_depress_CCT * 0.6901121) + (referral_CCT * 0.220459) + (histdep_CCT * 0.807517) + (histmh_CCT * -0.197546) + (consult_mean_CCT * 0.0.0041373) + (hist_depdrugs_CCT * -0.0660863) + (hist_somsym_CCT * 0.015973) + (CCT_agemths * -0.0003042) + (psych_other_CCT * -0.0130987)))) |
| **Age 21/22 YPA quest (*n* = 1,298)** | Depression | gen YPA_dep_SMFQ = (exp(-2.036359 + (curr_depdrugs_YPA * 0.7299155) + (hist_depdrugs_YPA * 0.6132707) + (referral_YPA * 0.5390764) + (hist_depress_YPA * 0.4207832) + (hist_depsymp_YPA * 0.1804129) + (curr_anxdrugs_YPA * 0.122329) + (curr_depsymp_YPA * 0.033265))) / (1 + (exp(-2.036359 + (curr_depdrugs_YPA * 0.7299155) + (hist_depdrugs_YPA * 0.6132707) + (referral_YPA * 0.5390764) + (hist_depress_YPA * 0.4207832) + (hist_depsymp_YPA * 0.1804129) + (curr_anxdrugs_YPA * 0.122329) + (curr_depsymp_YPA * 0.033265)))) |
| **Age 22/23 YPB quest (*n* = 1,325)** | Depression | gen YPB_dep_SMFQ = (exp(-5.804938 + (hist_depdrugs_YPB * 0.6876923) + (curr_depdrugs_YPB * 0.8200612) + (hist_depsymp_YPB * 0.5401723) + (curr_depress_YPB * 0.6056239) + (hist_anxsymp_YPB * -0.6341232) + (selfharm_YPB * 0.5377375) + (YPB_agemths * 0.0136081) + (curr_depsymp_YPB * 0.502017) + (referral_YPB * 0.2895768) + (asd_YPB * 0.7015883) + (hist_phob_YPB * 0.4473284) + (hist_depress_YPB * 0.0714968))) / (1 + (exp(-5.804938 + (hist_depdrugs_YPB * 0.6876923) + (curr_depdrugs_YPB * 0.8200612) + (hist_depsymp_YPB * 0.5401723) + (curr_depress_YPB * 0.6056239) + (hist_anxsymp_YPB * -0.6341232) + (selfharm_YPB * 0.5377375) + (YPB_agemths * 0.0136081) + (curr_depsymp_YPB * 0.502017) + (referral_YPB * 0.2895768) + (asd_YPB * 0.7015883) + (hist_phob_YPB * 0.4473284) + (hist_depress_YPB * 0.0714968)))) |

*Table S15:* In-sample and out-of-sample deviance ratios for common mental disorders (CMDs) or depression at each time-point. Deviance ratios are taken from logistic cross-validation lasso models. The full models are based on the set of all the primary care variables described in table S3. The 'diagnosis only' models contain just the relevant 'current' diagnosis variables (for depression models, this is just current depression diagnosis; for CMD models, this is current depression, anxiety and phobia diagnosis). The 'diagnosis, symptoms and treatment' models contain the relevant 'current' diagnosis, symptoms or treatment variables (for depression models, this is current depression diagnosis, current depressive symptoms and current antidepressant use; for CMD models, this is current depression, anxiety and phobia diagnosis, current depressive and anxiety symptoms, and current antidepressant and anti-anxiety medications).

| **Time-point (*n*)** | **Diagnosis** | **Lasso model** | **Sample** | **Deviance ratio** |
|---|---|---|---|---|
| **Age 15/16 TF3 clinic (*n* = 3,663)** | Depression | Full | Training | 9.9% |
| | | | Validation | -1.3% |
| | | Diagnosis only | Training | 0% |
| | | | Validation | -0.9% |
| | | Diagnosis, symptoms and treatment | Training | 0% |
| | | | Validation | -0.9% |
| | CMD | Full | Training | 8.4% |
| | | | Validation | 2.9% |
| | | Diagnosis only | Training | 2% |
| | | | Validation | -1.2% |
| | | Diagnosis, symptoms and treatment | Training | 1.6% |
| | | | Validation | -1.5% |
| **Age 16/17 CCS questionnaire (*n* = 3,213)** | Depression | Full | Training | 8.3% |
| | | | Validation | 4.3% |
| | | Diagnosis only | Training | 0.4% |
| | | | Validation | 0.3% |
| | | Diagnosis, symptoms and treatment | Training | 0.8% |
| | | | Validation | 1.8% |
| **Age 17/18 TF4 clinic (*n* = 3,084)** | Depression | Full | Training | 14.6% |
| | | | Validation | 7.8% |
| | | Diagnosis only | Training | 2.4% |
| | | | Validation | 1.1% |
| | | Diagnosis, symptoms and treatment | Training | 7.6% |
| | | | Validation | 6.9% |
| | CMD | Full | Training | 9.2% |
| | | | Validation | 7.8% |
| | | Diagnosis only | Training | 3.8% |
| | | | Validation | 4.9% |
| | | Diagnosis, symptoms and treatment | Training | 3.8% |
| | | | Validation | 5.2% |
| **Age 18/19 CCT questionnaire (*n* = 1,982)** | Depression | Full | Training | 9% |
| | | | Validation | 8% |
| | | Diagnosis only | Training | 1.3% |
| | | | Validation | 0.8% |

| | | | | |
|---|---|---|---|---|
| | | Diagnosis, symptoms and treatment | Training | 3.2% |
| | | | Validation | 5.3% |
| **Age 21/22 YPA questionnaire (*n* = 1,298)** | Depression | Full | Training | 11.7% |
| | | | Validation | 12.6% |
| | | Diagnosis only | Training | 3.3% |
| | | | Validation | 1.6% |
| | | Diagnosis, symptoms and treatment | Training | 8.8% |
| | | | Validation | 10% |
| **Age 22/23 YPB questionnaire (*n* = 1,325)** | Depression | Full | Training | 13.4% |
| | | | Validation | 9.1% |
| | | Diagnosis only | Training | 3.5% |
| | | | Validation | 2.7% |
| | | Diagnosis, symptoms and treatment | Training | 7.9% |
| | | | Validation | 5% |

*Table S16:* Sensitivities and specificities from the 40% validation sample, assessing how well the lasso model predicts common mental disorders (CMDs) or depression in ALSPAC across each of the time-points. Sensitivities and specificities are also given for the three 'CMD/depression' definitions using only primary care diagnosis, symptoms and/or treatment data (all using the same 40% validation sample). In these analyses we are treating the ALSPAC data as the reference standard. Note that for disclosure control purposes, statistics calculated where at least one cell in the cross-tabulation has a value <5 have been suppressed and replaced with a '<' or '>' summary statistic.

| | **Sensitivity** | **Specificity** |
|---|---|---|
| **Age 15/16 TF3 clinic depression – DAWBA (*n*=1,465; *n* with ALSPAC depression = 19)** | | |
| **Lasso prediction model** | 0% (0; 17.6)[a] | 100% (99.7; 100)[a] |
| **Current diagnosis** | 0% (0; 17.6)[a] | >99% |
| **Current diagnosis, treated** | 0% (0; 17.6)[a] | 100% (99.7; 100)[a] |
| **Current diagnosis or symptoms or treatment** | <10% | >99% |
| **Age 15/16 TF3 clinic CMD – DAWBA (*n*=1,465; *n* with ALSPAC CMD = 40)** | | |
| **Lasso prediction model** | 0% (0; 8.8)[a] | 100% (99.7; 100)[a] |
| **Current diagnosis** | <5% | >99% |
| **Current diagnosis, treated** | 0% (0; 8.8)[a] | 100% (99.7; 10)[a] |
| **Current diagnosis or symptoms or treatment** | <10% | >98% |
| **Age 16/17 CCS questionnaire depression – SMFQ (*n*=1,285; *n* with ALSPAC depression = 197)** | | |
| **Lasso prediction model** | 3.5% (1.4; 7.2) | 99% (98.2; 99.5) |
| **Current diagnosis** | <3% | >99% |
| **Current diagnosis, treated** | <3% | >99% |
| **Current diagnosis or symptoms or treatment** | 8.1% (4.7; 12.9) | 99% (98.2; 99.5) |
| **Age 17/18 TF4 clinic depression – CIS-R (*n*=1,234; *n* with ALSPAC depression = 100)** | | |
| **Lasso prediction model** | 6% (2.2; 12.6) | 99.1% (98.4; 99.6) |
| **Current diagnosis** | 5% (1.6; 11.3) | 99.3% (98.6; 99.7) |
| **Current diagnosis, treated** | <5% | >99% |
| **Current diagnosis or symptoms or treatment** | 26% (17.7; 35.7) | 97.3% (96.1; 98.1) |
| **Age 17/18 TF4 clinic CMD – CIS-R (*n*=1,234; *n* with ALSPAC CMD = 187)** | | |
| **Lasso prediction model** | 8% (4.6; 12.9) | 99.1% (98.4; 99.6) |
| **Current diagnosis** | 7% (3.8; 11.6) | 98.5% (97.5; 99.1) |
| **Current diagnosis, treated** | 3.7% (1.5; 7.6) | 99.4% (98.8; 99.8) |
| **Current diagnosis or symptoms or treatment** | 23% (17.2; 29.7) | 96.4% (95.1; 97.4) |
| **Age 18/19 CCT questionnaire depression – SMFQ (*n*=793; *n* with ALSPAC depression = 152)** | | |
| **Lasso prediction model** | 13.2% (8.2; 19.6) | 98.1% (96.8; 99) |
| **Current diagnosis** | 3.9% (1.5; 8.4) | 99.2% (98.2; 99.7) |
| **Current diagnosis, treated** | <5% | >99% |
| **Current diagnosis or symptoms or treatment** | 22.4% (16; 29.8) | 97.7% (96.2; 98.7) |

| | | |
|---|---|---|
| **Age 21/22 YPA questionnaire depression – SMFQ (*n*=519; *n* with ALSPAC depression = 82)** | | |
| **Lasso prediction model** | <10% | >99% |
| **Current diagnosis** | 7.3% (2.7; 15.2) | 98.6% (97) |
| **Current diagnosis, treated** | 6.1% (2; 13.7) | 98.9% (97.4; 99.6) |
| **Current diagnosis or symptoms or treatment** | 36.6% (26.2; 48) | 93.6% (90.9; 95.7) |
| **Age 22/23 YPB questionnaire depression – SMFQ (*n*=530; *n* with ALSPAC depression = 92)** | | |
| **Lasso prediction model** | 16.3% (9.4; 25.5) | 98.2% (96.4; 99.2) |
| **Current diagnosis** | <10% | >99% |
| **Current diagnosis, treated** | <10% | >99% |
| **Current diagnosis or symptoms or treatment** | 25% (16.6; 35.1) | 93.8% (91.2; 95.9) |

[a] One-sided 97.5% confidence interval.

*DAWBA:* Development and Well-Being Assessment; *SMFQ*: Short Mood and Feelings Questionnaire; *CIS-R:* Clinical Interview Schedule – Revised.

*Table S17:* Examining the association between primary care depression and common mental disorder (CMD) diagnoses and having ALSPAC data at each time point. For each time point, we present three sets of analyses: i) unadjusted models exploring the univariable associations between having ALSPAC data and both depression and CMDs; ii) models adjusting for sex; and iii) models adjusting for both sex and maternal education (a proxy for SEP; operationalised as a binary variable with the categories 'CSE/Vocational/O level' vs 'A level/Degree'). All results are odds ratios with 95% confidence intervals in brackets.

| | **Depression** | **CMDs** |
|---|---|---|
| **Age 15/16 TF3 clinic** | | |
| *Unadjusted model (n = 8,565)* | 1.17 (0.42; 3.23) | 1.2 (0.71; 2.04) |
| *Adjusted for sex (n = 8,565)* | 1.16 (0.42; 3.21) | 1.14 (0.67; 1.93) |
| *Adjusted for sex & maternal education (n = 7,342)* | 1.38 (0.41; 4.66) | 1.05 (0.58; 1.89) |
| **Age 16/17 CCS questionnaire** | | |
| *Unadjusted model (n = 8,555)* | 1.31 (0.61; 2.1) | 1.44 (0.92; 2.23) |
| *Adjusted for sex (n = 8,555)* | 1 (0.53; 1.86) | 1.28 (0.82; 2.01) |
| *Adjusted for sex & maternal education (n = 7,335)* | 1.09 (0.55; 2.13) | 1.18 (0.72; 1.93) |
| **Age 17/18 TF4 clinic** | | |
| *Unadjusted model (n = 8,045)* | 1.07 (0.67; 1.71) | 1.21 (0.86; 1.71) |
| *Adjusted for sex (n = 8,045)* | 0.93 (0.58; 1.48) | 1.07 (0.75; 1.52) |
| *Adjusted for sex & maternal education (n = 6,786)* | 0.99 (0.59; 1.66) | 1.04 (0.71; 1.54) |
| **Age 18/19 CCT questionnaire** | | |
| *Unadjusted model (n = 8,384)* | 0.99 (0.63; 1.57) | 1.32 (0.95; 1.82) |
| *Adjusted for sex (n = 8,384)* | 0.76 (0.48; 1.21) | 1.04 (0.75; 1.44) |
| *Adjusted for sex & maternal education (n = 7,066)* | 0.92 (0.56; 1.5) | 1.12 (0.79; 1.59) |
| **Age 21/22 YPA questionnaire** | | |
| *Unadjusted model (n = 6,353)* | 1.29 (0.88; 1.91) | 1.59 (1.17; 2.14) |
| *Adjusted for sex (n = 6,353)* | 1.06 (0.72; 1.57) | 1.33 (0.98; 1.81) |
| *Adjusted for sex & maternal education (n = 5,300)* | 1.12 (0.74; 1.7) | 1.38 (1; 1.91) |
| **Age 22/23 YPB questionnaire** | | |
| *Unadjusted model (n = 5,660)* | 1.22 (0.8; 1.86) | 1.83 (1.36; 2.47) |
| *Adjusted for sex (n = 5,660)* | 0.94 (0.61; 1.45) | 1.5 (1.1; 2.03) |
| *Adjusted for sex & maternal education (n = 4,668)* | 1.1 (0.69; 1.75) | 1.65 (1.82; 2.3) |
